# Air pollution, sociodemographic and health conditions effects on COVID-19 mortality in Colombia: an ecological study

**DOI:** 10.1101/2020.07.22.20159293

**Authors:** Laura A. Rodriguez-Villamizar, Luis Carlos Belalcazar-Ceron, Julián Alfredo Fernández-Niño, Diana Marcela Marín-Pineda, Oscar Alberto Rojas-Sánchez, Lizbeth Alexandra Acuña-Merchán, Nathaly Ramirez-Garcia, Sonia Cecilia Mangones-Matos, Jorge Mario Vargas-Gonzalez, Julián Herrera-Torres, Dayana Milena Agudelo-Castañeda, Juan Gabriel Piñeros Jiménez, Néstor Y. Rojas-Roa, Victor Mauricio Herrera-Galindo

**Author notes:** Correspondence Laura A. Rodriguez-Villamizar, Department of Public Health, Universidad Industrial de Santander, Bucaramanga, Colombia Carrera 32 29-31 Of. 301 Facultad de Salud.

## Abstract

**Objective:** To determine the association between chronic exposure to fine particulate matter (PM_2.5_), sociodemographic aspects, and health conditions and COVID-19 mortality in Colombia.

**Methods:** Ecological study using data at the municipality level, as units of analysis. COVID-19 data were obtained from official reports up to and including July 17th, 2020. PM_2.5_ long-term exposure was defined as the 2014-2018 average of the estimated concentrations at municipalities obtained from the Copernicus Atmospheric Monitoring Service Reanalysis (CAMSRA) model. We fit a logit-negative binomial hurdle model for the mortality rate adjusting for sociodemographic and health conditions.

**Results:** Estimated mortality rate ratios (MRR) for long-term average PM_2.5_ were not statistically significant in either of the two components of the hurdle model (i.e., the likelihood of reporting at least one death or the count of fatal cases). We found that having 10% or more of the population over 65 years of age (MRR=3.91 95%CI 2.24-6.81), the poverty index (MRR=1.03 95%CI 1.01-1.05), and the prevalence of hypertension over 6% (MRR=1.32 95%CI1.03-1.68) are the main factors associated with death rate at the municipality level. Having a higher hospital beds capacity is inversely correlated to mortality.

**Conclusions:** There was no evidence of an association between long-term exposure to PM_2.5_ and mortality rate at the municipality level in Colombia. Demographics, health system capacity, and social conditions did have evidence of an ecological effect on COVID-19 mortality.

## Introduction

The SARS-CoV-2 is a new coronavirus, responsible for the human coronavirus disease 2019 (COVID-19) initially reported in Wuhan, China in December 2019. The rapid global spread of COVID-19 made the World Organization of Health (WHO) to declare it a public health emergency of international concern (1). Up to and including July 20th 2020, 14,530,563 cases and 606,741 deaths have been reported in 188 countries (2). Approximately 80% of COVID-19 confirmed cases reported mild to moderate disease and the average case fatality rate is 4.6%, with a wide variation across countries (3).

Efforts to determine modifiable factors that could exacerbate symptoms and increase the risk of COVID-19 mortality remain essential to guide public policies. Conditions such as age above 65 years and underlying chronic diseases including diabetes, hypertension, cardiovascular disease, chronic lung disease, kidney failure, and cancer increase the risk of mortality (4-8). On the other hand, environmental factors have been less explored, despite the recent evidence of potential COVID-19 transmission through the air (9, 10). Recent studies suggest that chronic air pollution exposure, mainly to nitrogen dioxide (NO_2_), fine particulate matter (PM_2.5_) and sulphur dioxide (SO_2_), increase the COVID-19 mortality risk by 11.2% (CI95%: 3.4%-19.5%), 15% (CI95%:5% - 25%) and 17.2% (CI95%:0.5%-36.9%), respectively (11-13).

The dynamics of the pandemic have had geographic transitions, from China to Europe and then to America, with a growing impact in Latin America and the Caribbean (14, 15). In developing countries of Latin America, average concentrations of air pollutants are usually higher than levels reported in high-income countries of Europe and North America, especially for PM_2.5_ (16). Despite the known effect of chronic exposure to air pollution on the burden of cardiovascular and respiratory diseases (17, 18), its potential effect on COVID-19 mortality has not been fully elucidated, particularly in low-and-middle income countries. The objective of the study was to determine the association of chronic exposure to PM_2.5_, with COVID-19 mortality in Colombia, using an ecological approach.

## Methods

### Study Population

Colombia is a country located in the extreme north of South America, made up of 32 departments, 1,122 municipalities and a total population estimated by 2020 of 50,372,424 inhabitants according to the National Administrative Department of Statistics (DANE, for its initials in Spanish) (19). Half of the population are women (51.2%), 77.1% of people live in urban areas and 68.2% of Colombians are between 15 and 64 years old.

The first case of COVID-19 was confirmed on March 6th 2020 in Bogotá, the capital district. After nearly four months of the epidemic, there have been 182,140 confirmed cases of COVID-19 in 772 municipalities of Colombia up to and including July 17^th^, according to the National Institute for Health (INS, for its initials in Spanish). Most COVID-19 confirmed cases are male (53.6%) and 45,5% of cases are in the range between 20 and 39 years old. Deaths for COVID-19 reach 6,288 cases with a fatality proportion among confirmed cases of 3.4%. A total of 1,159,562 molecular tests have been conducted with a current test positivity of 17% (20).

### Data Sources

#### Air pollution data

Ninety-two out of 1122 municipalities measure air quality on a regular basis in Colombia (21). Large cities such as Bogota, Medellin, Bucaramanga, Cali, and Barranquilla have air quality monitoring networks. Medium-size and smaller cities perform periodic manual measurements that are not easily available. Existing surface measurements indicate that air quality problems are mainly associated with particulate matter (PM_10_ and PM_2.5_) as these pollutants regularly exceed air quality regulations, especially during the dry period, from January to April. Because of the scarcity of surface measurements in the country, we retrieved PM_2.5_ surface concentrations from the Copernicus Atmospheric Monitoring Service CAMS Reanalysis (CAMSRA) for this study. CAMSRA uses four-dimensional variational data assimilation techniques, combining satellite observations with a global scale atmospheric model to produce aerosol concentrations and mixing ratios of several gases at the surface and vertical gridded data (22, 23). CAMS Near Real-Time (CAMSNRT) is evaluated on a quarterly basis, and evaluation reports are available at the COPERNICUS website (24). Surface CAMSRA concentrations over Colombia for PM_2.5_ were downloaded using the ECMWF WebAPI and the Python script provided at this platform.Monthly average gridded data were retrieved at a 0.125-degree resolution from January 2014 to December 2018. PM_2.5_ concentrations were estimated at the centroid of each municipality by using a mathematical interpolation from the nearest four retrieved CAMSRA concentrations.

#### Population and socioeconomic data

Total population, population by age groups, and area (urban/rural) were retrieved at municipality level from the estimation of population 2020 based on the Colombian census DANE 2018.Cartographic information and maps were obtained from the DANE Geoportal public website(25) and the spatial data were created in ArcGIS 10.6.1® using the projection of Colombia in mode Custom Azimuth Equidistant and Datum WGS 1984. We used the multidimensional poverty index as a socioeconomic ecologic measure at the municipality level. The multidimensional poverty index is a national measure of poverty calculated at municipality level using data from the 2018 census of the population that includes 15 indicators grouped in five dimensions: home educational conditions, childhood and youth conditions, health, work, access to public services, and house conditions. The poverty index ranges from 0 to 100 with higher percentages meaning privation of more indicators and dimensions. The higher the index, the higher the socioeconomic deprivation (26).

#### Health data

For this study, data related to the number of confirmed cases and deaths for COVID-19 and the number of RT-PCR tests to confirm positive cases of infected people were taken from the official records of the National Institute for Health (INS). All 1,122 municipalities in Colombia report probable COVID-19 cases to the National Public Health Surveillance System (SIVIGILA, for its initials in Spanish) where cases are compiled, analyzed, and confirmed or discarded. Colombian official registries of COVID-19 cases and deaths are provided by INS and anonymized information is publicly available online (www.ins.gov.co). Database of cases includes case by case information of report date, diagnosis date, date of first symptoms, department and municipality of origin, age, sex, clinical condition, and death date for fatality cases. The INS also acts as the national laboratory of reference for laboratory tests. Information about the number of tests performed is updated daily by department and it is also available online at the INS website.

The crude period prevalence of arterial hypertension, diabetes mellitus, and chronic kidney disease was calculated for the 1,122 municipalities of Colombia. Prevalent cases were considered to be the people reported with the diagnosis for each disease by the Health Benefit Plan Management Entities (EAPB, for its initials in Spanish) to the High-Cost Account of the Ministry of Health and Social Protection between July 1st, 2018 and June 30th, 2019. According to the resolution 2463 of 2014 of the Ministry of Health and Social Protection of Colombia, the information of chronic kidney disease, hypertension, and diabetes mellitus patients should be reported annually with detailed case by case information including defined demographic and clinical indicators. Prior to calculations, these data were audited and filtered by the High-Cost Account for quality control. The denominator for the prevalence calculation was the estimated population by municipality projected by DANE on January 1st of 2019. Hospital beds capacity was measured as intermediate and intensive care beds per 100,000 inhabitants for each municipality, as a surrogate of the health system capacity. Data were obtained from the publicly available national registry of healthcare providers (Registro Especial de Prestadores de Servicios de Salud - REPS https://prestadores.minsalud.gov.co/habilitacion/).

#### Statistical analysis

The Colombian municipalities with at least one confirmed case of COVID-19 constituted the analytic sample. Population-time at risk was calculated as the total population multiplied by the number of days since the first symptom for the first confirmed case at each municipality with cases. The mortality proportion was calculated as the total confirmed deaths for COVID-19 divided by the total population, and the mortality rate was calculated using the population-time at risk as the denominator. The distribution of the deaths counts for the municipalities with at least one confirmed case of COVID-19 was graphically described, and its fit to a Poisson distribution was explored, using the variance test (VT) and the O_2_ test. Based on the mean (8.14) and variance (5,49) of the death counts the null hypothesis was rejected in both tests (p>0.01). The main source of overdispersion of the death counts of COVID-19 was given by the high heterogeneity of the counts (range: 0-1402), but also by the high presence of inflated zeros (58.5%).

Considering the high number of zeros and that from an epidemiological point of view the first death of COVID-19 represents a phase of the pandemic for one specific municipality, we decided to fit a hurdle model regression for the death counts. Hurdle models can be interpreted as two-part models integrated into one model. The first part is typically a binary response model (logit) and the second part is usually a truncated-at-zero count model (27). Consequently, the first component of the model lets us identify the variables associated with the positive outcomes (>0) that result from passing the zero hurdle (threshold), in this case defined as having at least one death. We fit a logit-negative binomial hurdle model as the counts over zero were still overdispersed. We use the population-time at risk as the “offset” variable in the regression models. These specifications let to estimate the effect of the independent variables on the expected logarithm of the mortality rate adjusted by person-time in each municipality.

The continuous long-term average of PM_2.5_ was used as the main independent variable in the hurdle model. In a sensitivity analysis, models were run using the PM_2.5_ average categorized and with restricted cubic splines using three knots. The Akaike criterion was calculated to compare the models. The effect of long-term PM_2.5_ was adjusted by the following confounding variables identified in the directed acyclic diagram (DAG, see supplementary material Figure S1) that were used as covariates: percentage of population 65 years or older, percentage of urban population, population density, poverty index, hospital beds capacity, number of COVID-19 tests at department level, and prevalence (percentage) of hypertension, diabetes, and chronic renal failure. We run the analysis clustered by department to account for potential correlation in municipalities within the same department. Secondary analyses were conducted excluding the capital district of Bogotá, which holds the highest count of deaths, excluding Medellin, the city among three capitals for which CAMSRA underestimates land-based concentrations (see supplementary material Figur S3), and excluding municipalities with less than 10 confirmed cases. All the analyses were run in STATA 15.

## Results

There were 182,140 confirmed cases and 6,288 confirmed deaths for COVID-19 in Colombia up to and including July 17^th^. COVID-19 cases were confirmed in 772 out of 1122 municipalities (68.8%) and deaths for COVID-19 were reported in 320 (41.5% of municipalities with cases). Table 1 summarizes the characteristics of the municipalities with COVID-19 confirmed cases. Mortality proportion for COVID-19 varies widely across municipalities with confirmed cases from 0 to 197.0 per 100,000; the mortality rate (using person-time at risk as the denominator) ranged between 0 and 38.4 per 1,000,000. Figure 1(a) presents the mortality rate for COVID-19 by municipality. The long-term average exposure to PM_2.5_ (2014-2018) in municipalities with COVID-19 confirmed cases was 20.0 µg/m^3^ with a range between 9.1 and 37.5 µg/m^3^. Figure 1(b) presents the long-term average exposure to PM_2.5_ at municipality level.

**Table 1.** Characteristics of the municipalities with COVID-19 cases in Colombia up to and including July 17^th^, 2020 (mean and SD)

| Variable | Total<br>772 municipalities | PM<sub>2.5</sub><20 µg/m<sup>3</sup><br>393 municipalities | PM<sub>2.5</sub>≥20 µg/m<sup>3</sup><br>379 municipalities |
| --- | --- | --- | --- |
| COVID-19 mortality rate (per 1,000,000) | 0.75 (2.23) | 0.75 (2.60) | 0.74 (1.76) |
| Time since symptoms in first case | 66.15 (37.58) | 62.48 (36.68) | 69.96 (38.18) |
| % Population 65 or older | 10.03 (3.64) | 9.89 (3.75) | 10.17 (3.53) |
| % Urban population | 49.26 (24.71) | 41.74 (22.53) | 57.07 (24.48) |
| Population density per Km<sup>2</sup> | 229.98 (899.66) | 116.38 (421.68) | 347.78 (1119.66) |
| Poverty index | 39.85 (17.73) | 42.56 (18.72) | 37.03 (16.19) |
| Hospital beds capacity (per 100,000) | 71.95 (78.22) | 70.78 (77.73) | 73.15 (78.81) |
| % Hypertension | 6.11 (3.74) | 5.68 (3.19) | 6.57 (4.19) |
| % Diabetes | 1.57 (1.42) | 1.43 (1.04) | 1.73 (1.71) |
| %Chronic renal failure | 0.94 (1.26) | 0.86 (1.00) | 1.04 (1.48) |

**Figure 1.**
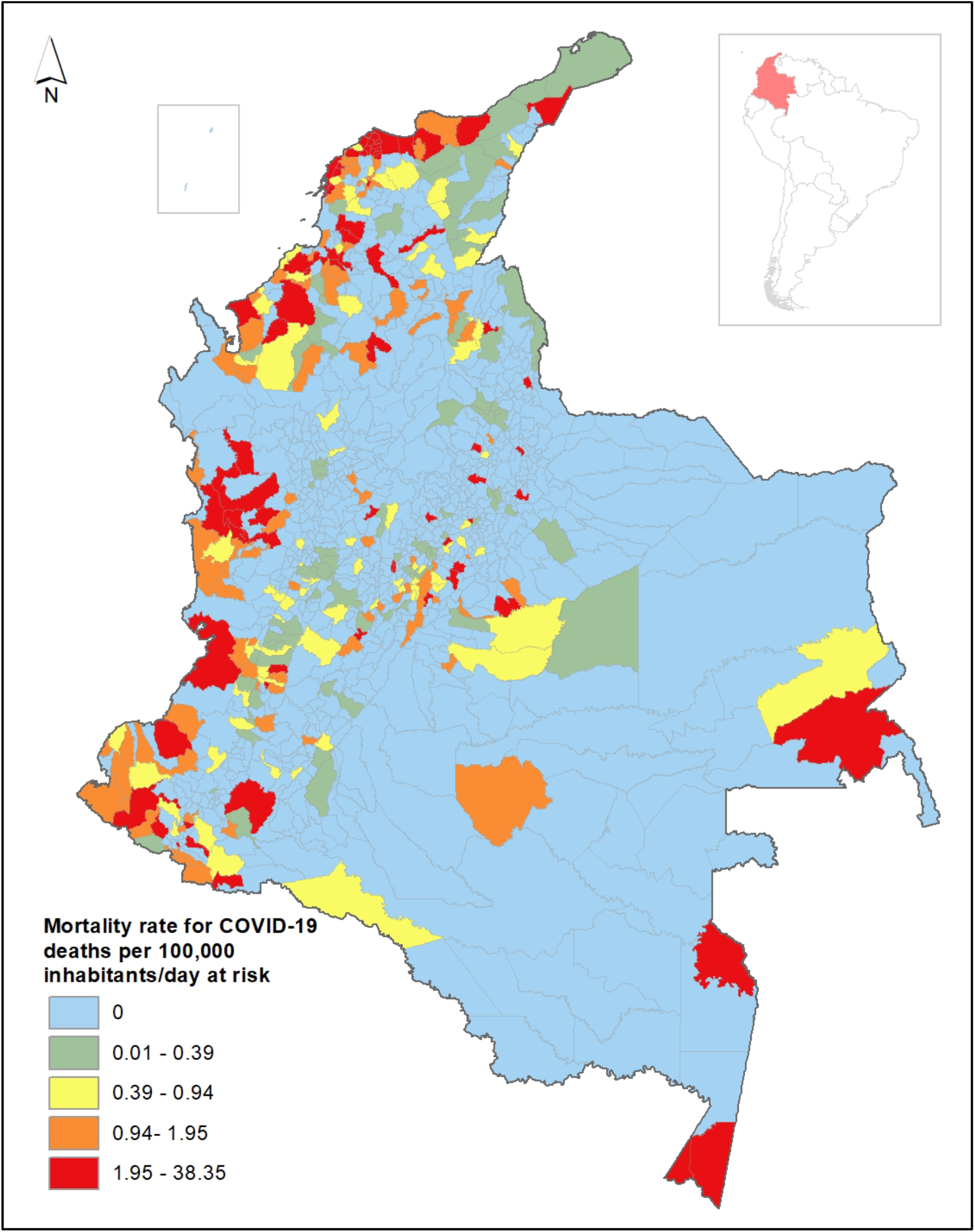
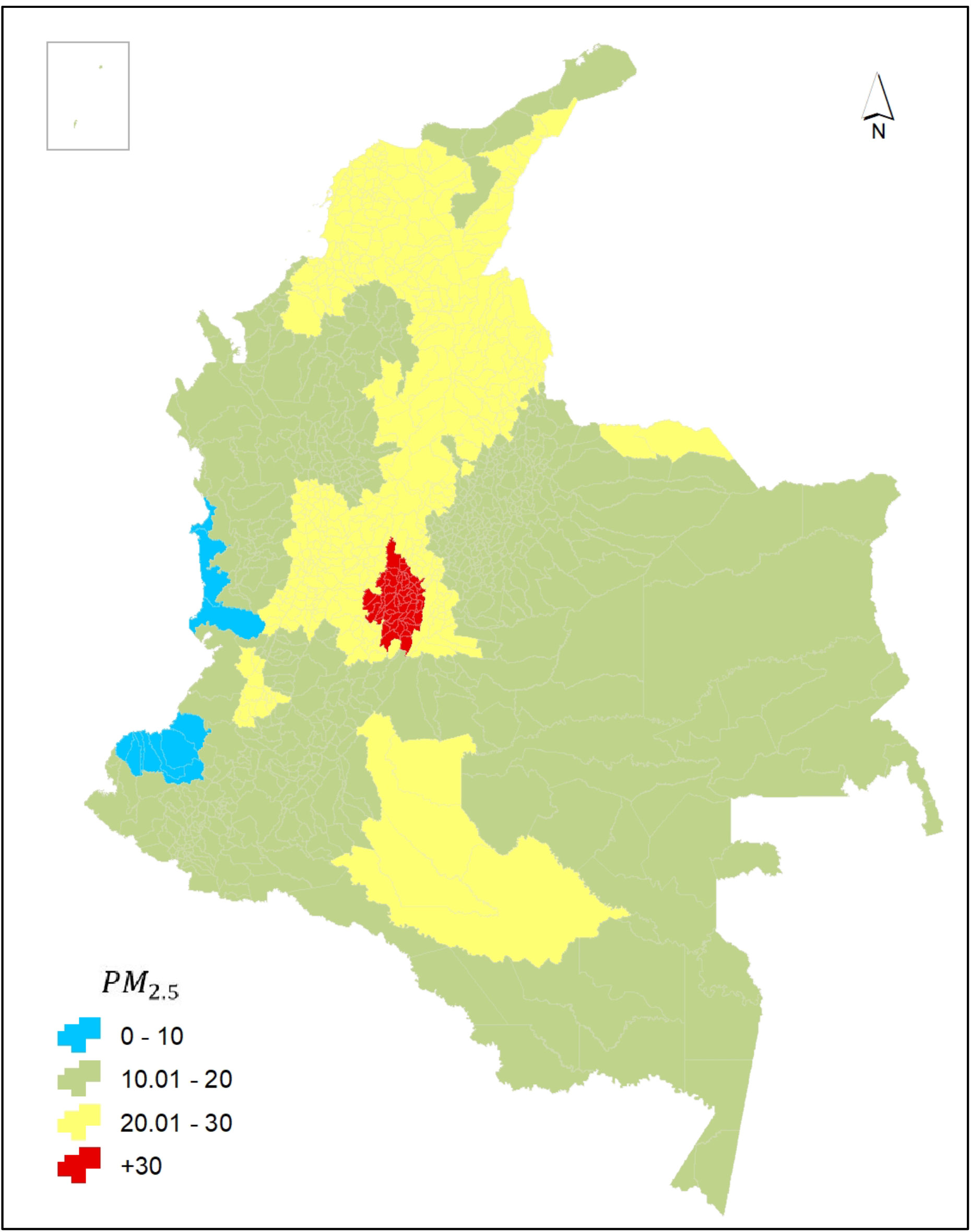
Mortality for COVID-19 and PM_2.5_ long-term average in Colombia at municipality level. (a) Mortality rate for COVID-19 by municipality in Colombia up to and including July 17^th^, 2020. (b) Long-term average of PM_2.5_ concentrations (2014-2018) in Colombia (in *μ*g/m^3^)

The geographic pattern of COVID-19 mortality proportion and long-term average of PM_2.5_ do not seem to have a good overlap at visual inspection as some municipalities with low levels of PM_2.5_ exhibit high mortality proportion. The regions with higher mortality proportion are located in the Atlantic and Pacific coasts, and the Amazonian region. Figure 2 shows the graphical inspection of the relation between the estimated long-term mean of PM_2.5_ and the logarithm of the COVID-19 mortality rate. The patterns did not follow a linear trend and increased log of mortality rates for COVID-19 are present at lowest levels of mean PM_2.5_. Using a binomial approach (having or no having deaths), the relation with mean PM_2.5_ did not follow a linear trend but a line with different inflection points (See Supplementary material Figure S2). Restricted cubic splines of PM_2.5_ with three knots identified those points to be 12.6, 19.3, and 26.6 µg/m^3^.

**Figure 2.**
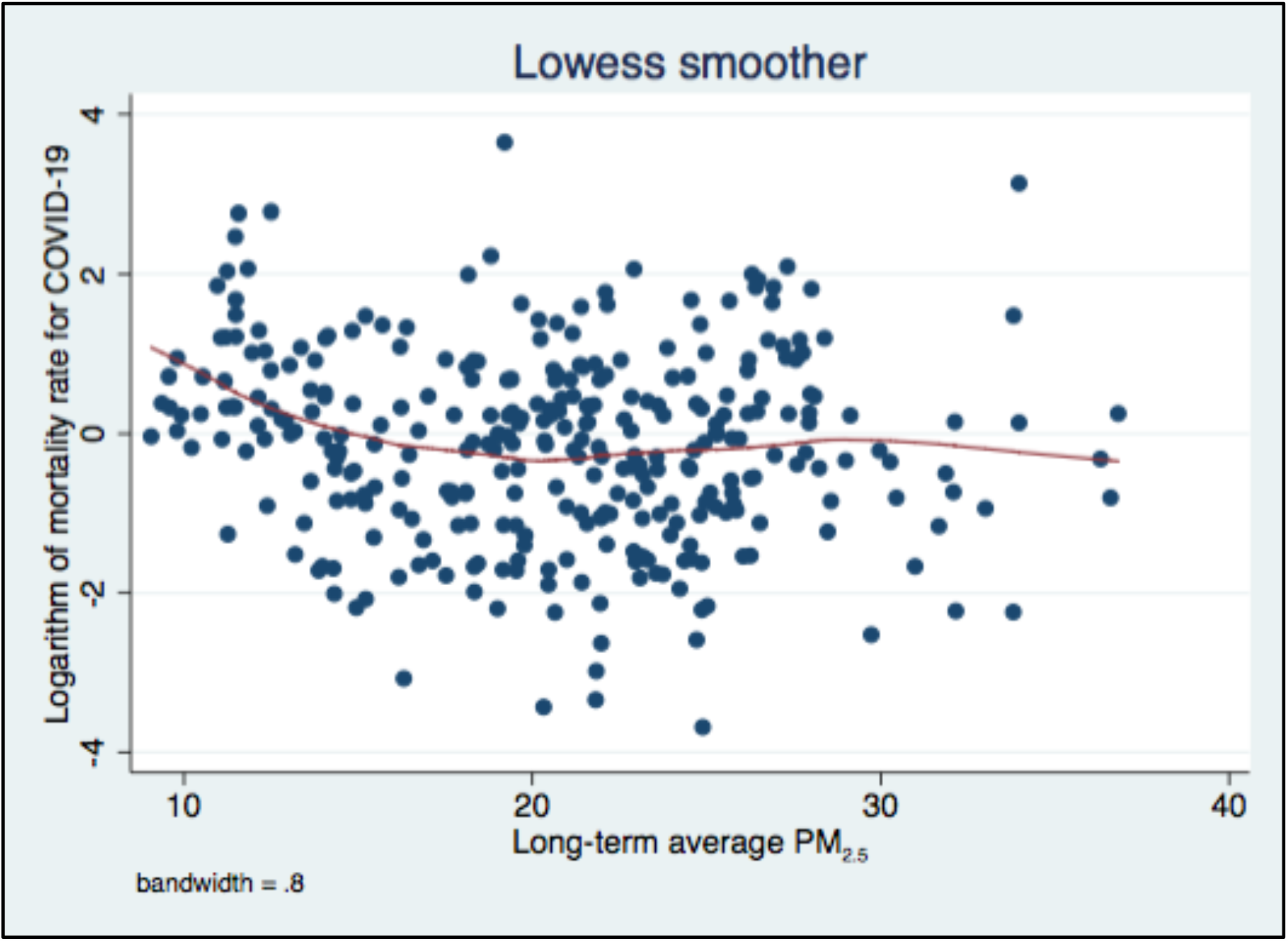
Relation between mortality rate for COVID-19 and PM_2.5_ in municipalities with COVID-19 confirmed cases in Colombia up to and including July 17^th^, 2020.

We present the results of our main analysis using hurdle models in Table 2. Estimated mortality rate ratios (MRR) for long-term average PM_2.5_ were not statistically significant in either of the two components of the model: the logit component modeling the change of no having deaths to have at least one death, and the negative binomial model of counts of deaths. In the logit component, we found that having 10% or more of the population over 65 years of age and the prevalence of hypertension over 6% are the main factors associted with death rate at municipality level while having a higher percentage of urban population and higher hospital beds capacity are negatively correlated to mortality. Once the municipality reaches at least one COVID-19 death, the main factors associated with the mortality rate are the percentage of urban population and the poverty index. In addition, an important cluster (department) effect was identified in the data (p-value=0.001).

**Table 2.** Mortality rate ratios in the main analysis using hurdle models for municipalities with COVID-19 cases in Colombia up to and including July 17^th^, 2020

| Variable | Component logit<br>(0-at least one death) |  |  | Component negative binomial<br>(counts of deaths>1) |  |  |
| --- | --- | --- | --- | --- | --- | --- |
|  | MRR | 95% CI | P-value | MRR | 95% CI | P-value |
| Long-term average PM<sub>2.5</sub> (µg/m<sup>3</sup>) | 1.00 | 0.92 - 1.08 | 0.973 | 1.00 | 0.95 - 1.06 | 0.747 |
| % Population 65 or older>10% | 3.91 | 2.24 – 6.81 | 0.000 | 0.51 | 0.23 – 1.14 | 0.100 |
| % Urban population | 0.96 | 0.94 – 0.98 | 0.000 | 1.02 | 1.01 – 1.03 | 0.000 |
| Population density (per Km<sup>2</sup>) | 0.99 | 0.99 – 1.00 | 0.129 | 1.00 | 0.99 – 1.00 | 0.033 |
| Poverty index | 0.99 | 0.97 – 1.02 | 0.928 | 1.03 | 1.01 - 1.05 | 0.001 |
| Hospital beds capacity (per 100,000) | 0.99 | 0.98 – 0.99 | 0.000 | 1.00 | 0.99 – 1.01 | 0.052 |
| % Hypertension |  |  |  |  |  |  |
| spline 1 (2.09-5.97) | 0.75 | 0.60 – 0.94 | 0.013 | 0.96 | 0.73 – 1.28 | 0.804 |
| spline 2 (5.97-10.37) | 1.32 | 1.03 – 1.68 | 0.026 | 1.00 | 0.70 – 1.45 | 0.960 |
| % Diabetes>4% | 0.29 | 0.01 – 9.97 | 0.495 | 1.74 | 0.43 – 6.95 | 0.434 |
| %Chronic renal failure>3% | 1.25 | 0.35 – 4.39 | 0.726 | 0.42 | 0.15 – 1.17 | 0.096 |
| Number of test at department level | 1.00 | 0.00 - 1.00 | 0.056 | 1.00 | 0.99 - 1.00 | 0.565 |

MRR: Mortality Rate Ratio; CI: Confidence Interval

We found that secondary analysis exhibits similar results to our main analysis in terms of no evidence of increased risk of COVID-19 mortality rate associated with increased long-term average of PM_2.5_ at municipality level (Table 3). Results similar to our main analysis were also consistent in sensitivity analysis using different approaches to model PM_2.5_ long-term average exposure (See supplementary material Table S1).

**Table 3.** Mortality rate ratios in secondary analysis using hurdle models for municipalities with COVID-19 cases in Colombia up to and including July 17^th^, 2020

| Long-term PM<sub>2.5</sub><br>($\mu\text{g}/\text{m}^3$) | Component logit*<br>(0 - at least one death) | | | Component negative binomial*<br>(counts of deaths > 1) | | |
| --- | --- | --- | --- | --- | --- | --- |
|  | MRR | 95% CI | P-value | MRR | 95% CI | P-value |
| Excluding Bogotá | 1.00 | 0.93 - 1.08 | 0.973 | 1.00 | 0.96 - 1.06 | 0.784 |
| Excluding Medellín | 1.00 | 0.93 - 1.08 | 0.973 | 1.01 | 0.95 - 1.06 | 0.759 |
| Excluding<br>municipalities with less<br>than 10 confirmed<br>COVID-19 cases | 1.02 | 0.96 - 1.07 | 0.494 | 1.03 | 0.96 - 1.09 | 0.347 |

MRR: Mortality Rate Ratio; CI: Confidence Interval
\*Adjusted for the percentage of population 65 years or older, percentage of urban population, population density, poverty index, hospital beds capacity, number of tests at department level, and prevalence of hypertension, diabetes, and chronic renal failure.

## Discussion

This is the first ecologic nation-wide study conducted in a developing country assessing the association between COVID-19 mortality and long-term exposure to PM_2.5_. Our results did not find evidence of an association between higher concentrations of PM_2.5_ and higher counts of deaths, controlling for nine socioeconomic and health indicators at municipality level. The effect of socioeconomic and health conditions in the municipalities, such as the proportion of the population over 65 years, the poverty index, and the prevalence of hypertension, did show evidence of increasing the risk of deaths for COVID-19, while the hospital’s capacity decreased that risk.

The first COVID-19 confirmed case was identified in Bogota, the capital district (20), where the highest number of cases have occured exceeding 45,000 by mid-July. The first cases identified in the capital district were related to returning flights from Europe which were also related to the first cases identified in other main capital cities including Medellin and Cali. Then, an additional infection source came from international cruises in the Port of Cartagena, where the epidemic spread to the Atlantic Coast. The epidemic in the Brazilian Amazonian region was the probable source of infection in Leticia, the capital of the Amazonas department, which is the municipality with the highest mortality proportion for COVID-19 in Colombia (195.02 per 100,000 inhabitants). Thus, the COVID-19 spread went from capital cities to small municipalities, where the mortality rates have been higher.

There was no evidence of an association between long-term average of PM_2.5_ and the mortality rate for COVID-19 in the crude or adjusted models. Our results contrast with studies conducted in China (12) and the United States (11), which found positive associations between PM_2.5_ and COVID-19 mortality after adjusting for four and 20 potential confounders, respectively. These results supported the hypothesis that the effect of long-term exposure to PM_2.5_ on COVID-19 mortality is largely mediated by comorbidities linked to chronic PM-related inflammation (28, 29). In this regard, it has been proposed that chronic exposure to PM_2.5_ causes alveolar ACE-2 receptor overexpression, that may increase viral load in patients exposed to pollutants (30). Our findings revealed a significant effect of ageing and poverty on COVID-19 mortality rate, factors related to failure in the mechanisms of acute immune humoral and cellular response at the individual level, and to higher burden of chronic disease and lower capability of the healthcare system to treat complicated cases of infection, at the municipal level. These findings might suggest that the chronic effect of ageing and poverty might have a stronger effect on COVID-19 complications and mortality in developing countries.

Another possible explanation to our findings is that long-term exposure to PM_2.5_ has less impact on biological susceptibility to COVID-19 complications and deaths compared to the effect of other air pollutants such as nitrogen dioxide (NO_2_). The stronger short-term effect of NO_2_ compared to other pollutants on respiratory and cardiovascular morbidity have been identified using multipollutant models in Colombia (31). A country-wide cross-sectional study in the United States using multipollutant models for the effect of PM_2.5_, NO_2_ and O_3_ found a strong positive association between NO_2_ and COVID-19 fatality and mortality but did not find significant associations with PM_2.5_ and O_3_ (13). The authors discussed that divergent results with the previous US nationwide study (11) are probably due to the use of multi-pollutant models and the adjustment for spatial trends, which might have confounded the findings. Unfortunately, we did not count on reliable NO_2_ and O_3_ long-term exposure estimations so we did not assess this effect in multi-pollutant models.

We found an independent and significant effect of the older age, the poverty index, and the prevalence of hypertension (over 6%) associated with COVID-19 mortality rate. Similar findings related to age and chronic diseases have been reported in several studies (4-8). In Italy Conticini et al. (28) discussed that important factors such as the age structure of the affected population, the great differences between the Italian regional health systems, the capacity of intensive care units in the region, and prevention policies adopted by the government have played a major role in the spread of and mortality for SARS-CoV-2, presumably more than long-term air pollution itself. The effect of poverty on COVID-19 mortality has been less described in the literature but it represents a major risk condition in developing countries probably related to unstable occupation and income, lower health literacy, and limited access to preventive health services (32, 33). A few recent ecological studies in the US at county level have reported a correlation between COVID-19 mortality rate and some social disparities such as poverty status and non-English speaking households and other ethnic minorities (34, 35).

Strengths of this study include the use of nationwide public official data at municipality level, adjustment for nine sociodemographic and health conditions using a hurdle model, and the use of average long-term estimations of PM_2.5_ exposure. The estimation of PM_2.5_ concentration in this study comes from the CAMSRA model, which has been evaluated using independent measurements available in different world regions at the ground level and in the tropospheric column. These evaluations show that CAMSRA successfully reproduces levels and trends of aerosols and gases (36). An evaluation of the performance of CAMSRA over the cities of Bogota, Medellin, and Bucaramanga for PM_2.5_, CO, and NO_2_ concentrations was conducted recently comparing measurements from the air quality monitoring networks with retrieved CAMSRA concentrations. Results showed that CAMSRA is able to reproduce PM_2.5_ levels and trends in these three cities, however, the model largely underestimates NO_2_ and CO concentrations (37) (also see figure S3 in the supplementary material). Although elevated levels of PM_2.5_ are observed in urban areas, PM_2.5_ distribution in Colombia shows that even medium-size and small municipalities have similar or even higher concentrations of PM_2.5_. This behavior coincides with the geographical distribution of aerosols reported in previous studies for Colombia (18, 38). These studies indicate that biomass burning is an important source of PM_2.5_ in Colombia and that both large and small cities are affected by this source. The PM_2.5_ geographical distribution and trends presented in this study (Figure 1b) coincides with results reported in all these studies.

Our study has some limitations. First, it was conceived as an ecological study whose nature precludes the extrapolation of inferences from the empirical evaluation of hypothesis based on clusters (i.e., municipalities) to the individual level, therefore, the absence of a relationship between long-term exposure to PM_2.5_ and mortality among patients diagnosed with SARS-CoV-2 should at best be regarded as provisional. Second, exposure was determined using a model-based approach, that even though captured the heterogeneity of concentration across municipalities had a moderate correlation to land-based measurements in a selected sample of some of the largest ones, which might have partially attenuated any underlying association between PM_2.5_ and mortality. Third, in the absence of reliable NO_2_ and O_3_ long-term exposure estimations, we could not incorporate them into the analysis or evaluate the independent association of PM_2.5_ and mortality in the context of multi-pollutant models. Fourth, mortality data reflect fatal cases among patients with a confirmatory diagnosis of the infection, excluding deaths among undiagnosed individuals (due to either low testing rates or unreliable test results) and those occurring outside of hospitals. Systematic differences in the capability of municipalities to comprehensively and correctly identify and register deaths attributable to the infection could have biased our estimate of effect. Although this issue could not be directly addressed in the analysis, adjusting for testing rates and hospital beds capacity should have partially corrected for differential readiness of municipal health systems to cope with the epidemic.

## Conclusions

There was no evidence of an association between long-term exposure to PM_2.5_ and mortality rate for COVID-19 at the municipality level in Colombia. Demographics, health system capacity, and social conditions did show an ecological effect on COVID-19 mortality.

## Data Availability

Data will be made available upon request

## Data Sharing statement

Data will be made available upon request

## Acknowledgement

The authors would like to thank Yurley Rojas for her contribution to the generation of maps and Gloria Ramos for updating the COVID-19 public data set for analysis.

